# Inferring Clinically Relevant Molecular Subtypes of Pancreatic Cancer from Routine Histopathology Using Deep Learning

**DOI:** 10.64898/2026.01.06.26343539

**Authors:** Abdul Rehman Akbar, Alejandro Levya, Ashwini Esnakula, Elshad Hasanov, Anne Noonan, Upender Manne, Vaibhav Sahai, Lingbin Meng, Susan Tsai, Anil Parwani, Wei Chen, Ashish Manne, Muhammad Khalid Khan Niazi

**Author notes:** The authors contributed equally to this work. The names are mentioned in alphabetical order. These authors jointly supervised this work (Ashish Manne, Muhammad Khalid Khan Niazi). Correspondence (A.R.A.).

## Abstract

**Background and aims:** Molecular subtyping of pancreatic ductal adenocarcinoma (PDAC) into basal-like and classical has established prognostic and predictive value. However, its use in clinical practice is limited by cost, turnaround time, and tissue requirements, thereby restricting its application in the management of PDAC. We introduce PanSubNet (PANcreatic SUBtyping NETwork), an interpretable deep learning framework that predicts therapy-relevant molecular subtypes directly from standard hematoxylin and eosin (H&E)-stained whole-slide images.

**Methods:** PanSubNet was developed using data from **1**,**055 patients** across two multi-institutional cohorts (PANCAN, n=846; TCGA, n=209) with paired histology and RNA sequencing data. Ground-truth labels were derived using the validated Moffitt 50-gene signature refined by GATA6 expression. The model employs dual-scale architecture that fuses cellular-level morphology with tissue-level architecture, leveraging attention mechanisms for multi-scale representation learning and transparent feature attribution.

**Results:** On internal validation within PANCAN using five-fold cross-validation, PanSubNet achieved mean area under the receiver operating characteristic curve (AUC) of **88.5%** in high-confidence cases, with balanced sensitivity and specificity. External validation on the independent TCGA cohort without fine-tuning demonstrated robust generalizability (AUC **84.0%**). PanSubNet preserved and, in metastatic disease, strengthened prognostic stratification compared to RNA-seq–based labels. Prediction uncertainty linked to intermediate transcriptional states, not classification noise. Model predictions are aligned with established transcriptomic programs, differentiation markers, and DNA damage repair signatures.

**Conclusions:** By enabling rapid, cost-effective molecular stratification from routine H&E-stained slides, PanSubNet offers a clinically deployable and interpretable tool for genetic subtyping. We are gathering data from two institutions to validate and assess real-world performance, supporting integration into digital pathology workflows and advancing precision oncology for PDAC.

## Introduction

Pancreatic ductal adenocarcinoma (PDAC) is among the most lethal human malignancies. The overall 5-year survival for all-stage PDAC is approximately 11%, rising only to 42% for localized tumors and plummeting to 3% for metastatic tumors [1]. These outcomes stand in stark contrast to survival rates exceeding 90% for breast and prostate cancers, and approximately 65% for colorectal cancers. Despite this disproportionate mortality, systemic therapy selection in PDAC remains predominantly empiric. Across disease stages, treatment decisions are typically limited to two first-line regimens, (modified) FOLFIRINOX (irinotecan, oxaliplatin, and 5-fluorouracil) and gemcitabine/nab-paclitaxel (Gem-NP), with selection driven primarily by the patient fitness (performance status and age) rather than tumor biology [2–5]. This reflects both the absence of definitive comparative trials in advanced disease and the lack of clear superiority of either regimen in randomized studies of early-stage disease. Therapeutic progress over the past decade has been modest. Although the NAPOLI-3 trial showed improved outcomes with NALIRIFOX (nanoliposomal irinotecan, oxaliplatin, and 5-fluorouracil) compared to Gem-NP, survival metrics were comparable to those of FOLFIRINOX reported in the PRODIGE trial, underscoring the incremental rather than transformative gains achieved by irinotecan modification [6].

Routinely used biomarkers, including carbohydrate antigen 19-9 (CA 19-9), tissue mutation profiling, and cell-free DNA testing (liquid biopsy), provide limited prognostic insight and have little influence on treatment selection. Platinum-containing (cisplatin or oxaliplatin) regimens are preferentially considered for patients harboring select germline DNA damage repair alterations (e.g., *BRCA1/2, PALB2, ATM*, and *RAD50*); however, such alterations are rare, occurring in fewer than 10% of PDACs and up to 17% of familial cases [7]. Consequently, the vast majority of patients lack actionable biologic stratification at treatment initiation.

The clinical consequences of empiric therapy selection are most pronounced in settings with narrow therapeutic latitude. In potentially resectable tumors, ineffective initial treatment may permit progression to unresectable or locally advanced states, while in metastatic disease, early toxicity or rapid clinical decline may preclude subsequent lines of therapy. These constraints emphasize the need for earlier and more precise biologic stratification to inform treatment selection in PDAC.

Multiple molecular classification strategies, including mutational, proteomic, and transcriptomic signatures, have been proposed but remain largely confined to research [8–10]. Among them, transcriptomic subtyping into classical and basal-like phenotypes proposed by Moffitt et al., has demonstrated consistent prognostic (clinical outcomes) and predictive (treatment response) relevance across cohorts [11]. The Purity Independent Subtyping of Tumors (PurIST) classifier developed by Rashid et al. provides a simplified, gene-pair-based implementation of classical and basal-like subtyping; however, its classifications are not fully concordant with the original Moffitt transcriptomic framework [12]. Classical PDAC is associated with superior outcomes and greater sensitivity to FOLFIRINOX, while basal-like PDAC portends poor prognosis and limited benefit from this regimen [11–22]. In a therapeutic landscape lacking biologically guided treatment algorithms, transcriptomic subtyping could, in principle, support treatment selection and prognostication. In practice, widespread clinical adoption of transcriptomic subtyping remains constrained by cost, turnaround times, limited global availability of RNA sequencing, and frequent tissue inadequacy, particularly in small diagnostic biopsies. Collectively, these constraints underscored the need for alternative strategies that can derive biologically meaningful feedback subtypes from routinely available clinical material.

Deep learning applied to routine hematoxylin and eosin (H&E)-stained whole-slide images (WSIs) offers a scalable digital pathology approach to inferring molecular insights. H&E slides are ubiquitously available from standard diagnostic workflows, and various deep learning-based studies have demonstrated a remarkable ability to infer molecular features, including gene expression signatures and mutations, directly from histology in several cancers [23–32]. By leveraging existing digital pathology infrastructure, such approaches enable rapid and cost-efficient molecular stratification compatible with routine clinical workflows, supporting broader clinical implementation in PDAC management.

Building on this foundation, we present PanSubNet (PANcreatic SUBtyping NETwork), a WSI–based deep learning framework for PDAC molecular subtyping. PanSubNet utilizes 1,055 patients with H&E WSIs and RNA-seq–derived labels using Moffitt 50-gene signature–based basal/classical scores refined by GATA6 expression as ground truth [33, 34]. The model operates at both cellular and tissue scales, integrating cell-level features with architectural context to generate slide-level subtype predictions (see Figure 1). By demonstrating that biologically meaningful PDAC subtype information, traditionally derived from transcriptomic profiling, can be inferred directly from routine histopathology, PanSubNet establishes WSIs as a scalable framework for biological stratification. In this study, PDAC molecular subtyping serves as a clinically relevant use case illustrating how WSI-based models extract tumor intrinsic biology from standard diagnostic materials, complementing existing molecular assays and remaining compatible with existing clinical workflows.

**Figure 1.**
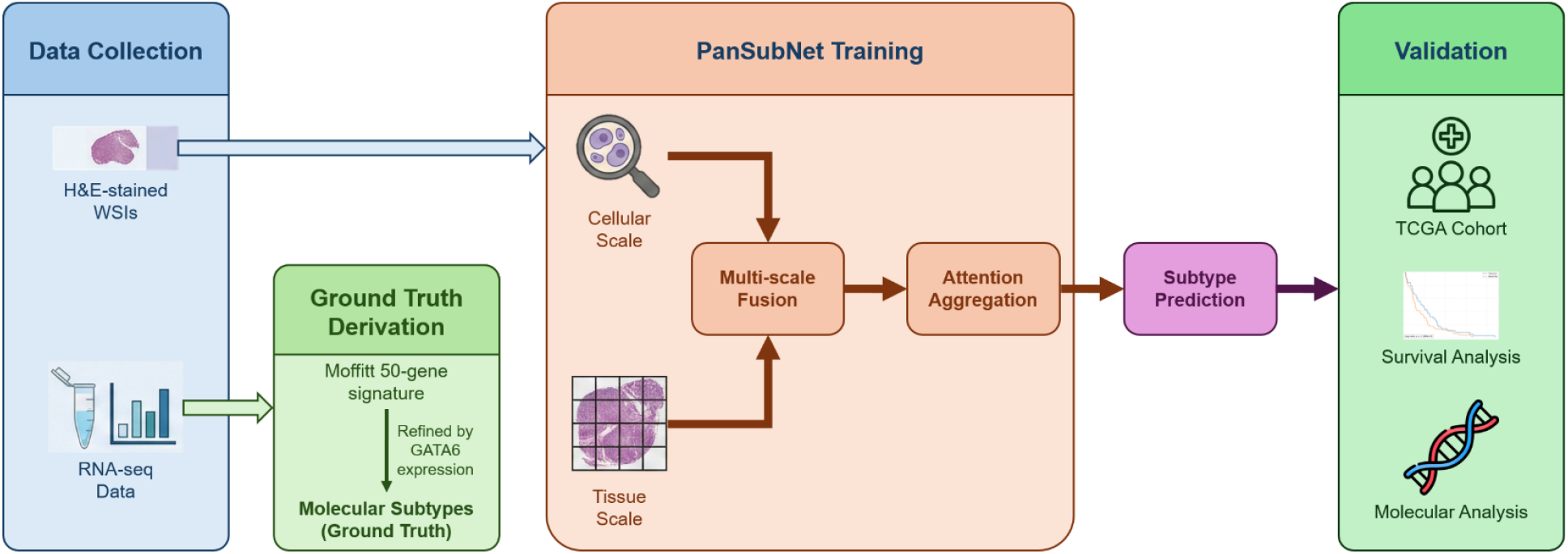
Schematic overview of the PanSubNet study workflow. The study methodology integrates paired H&E whole-slide images (WSIs) with transcriptomic data. Ground truth molecular subtypes are established using the Moffitt 50-gene signature, refined by GATA6 expression levels. The PanSubNet deep learning architecture employs a dual-scale approach to analyze the H&E slides: (1) a cellular scale utilizing CellViT++ to extract cell-level features and spatial context, and (2) a tissue scale utilizing UNI2-h to encode global patch-level features. These multi-scale representations are synthesized via a fusion and attention mechanism to generate a slide-level subtype prediction. The clinical and biological relevance of the model is validated through Kaplan-Meier survival analysis, assessment of DNA damage repair (DDR) gene associations, and functional pathway enrichment analysis.

Recent advances in computational biology have demonstrated the feasibility of molecular features from WSIs using deep learning models [32, 35]. Prior approaches have primarily focused on learning associations between patch-level or slide-level image features molecular levels, establishing proof of concept for histology-based molecular inference across multiple cancer types. However, many existing methods treat molecular prediction as a hard classification problem and rely predominantly on regional correlations, with limited explicit modelling of cellular morphology, spatial organization, or biological uncertainty. Here, we address these considerations by developing a framework that integrates multi-scale histologic context for molecular inference from routine WSIs.

## Materials and Methods

### Datasets and RNA-sequencing preprocessing

Our study comprised 1,055 patients across two cohorts: the PANCAN cohort (n = 846) and the TCGA-PAAD cohort (n = 209). All patients had diagnostic formalin-fixed, paraffin-embedded (FFPE) whole-slide images (WSIs) scanned at ×40 magnification. From this total, 792 patients (614 from PANCAN, 183 from TCGA) had paired H&E WSIs and bulk RNA sequencing data, which formed the dataset for downstream analysis.

For TCGA-PAAD, raw RNA-seq data were obtained through the Genomic Data Commons (GDC) manifest system. Expression values were downloaded as gene-level count matrices and converted to transcripts per million (TPM) using GENCODE gene models. Gene identifiers (ENSEMBL) were mapped to standard HUGO gene symbols using the corresponding GTF annotation [36, 37]. Expression values were extracted specifically for the 50-gene Moffitt classical/basal signature, and TPM values were z-scored across samples to normalize gene-level variation.

For the PANCAN cohort, raw FASTQ files were downloaded and processed following a consistent, uniform RNA-seq pipeline. Firstly, QC was performed on raw FASTQ files to assess GC content, sequence duplication, read saturation, and potential contamination. Secondly, transcript quantification was performed using Salmon (v1.10.2) in quasi-mapping mode with the appropriate Salmon index constructed from the same GENCODE reference used for TCGA [38]. Thirdly, transcript counts across all transcript isoforms were aggregated into gene-level abundances. Resulting TPM matrices were cross-checked to ensure complete coverage of all 50 Moffitt genes.

For both cohorts, single-sample gene-set enrichment analysis (ssGSEA) was used to compute classical and basal-like enrichment scores based on the Moffitt 50-gene signature. For each sample, genes were ranked by TPM; ssGSEA enrichment was computed separately for classical and basal gene sets; and a molecular subtype score was calculated as:

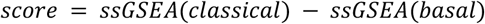

Positive values indicate classical-like expression; negative values indicate basal-like expression.

Subtype scores were then z-scored across all 792 patients to quantify the confidence of each classification. We defined:

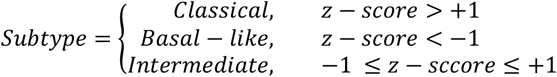

This thresholding reproduces the expected ~65/35 classical/basal distribution reported in prior studies and yields robust subtype assignments suitable for supervised learning.

Across the full cohort, 238 patients met high-confidence criteria (114 basal-like, 124 classical), and 554 patients fell into the intermediate range. High-confidence cases were used for all supervised training, internal validation, and external testing. Intermediate cases were excluded from model training but retained as a biological reference group for other downstream analysis tasks.

GATA6 is a well-established marker of the classical lineage and distinguishes classical from basal tumors in multiple transcriptomic frameworks. To refine ambiguous intermediate cases, GATA6 TPM values were stratified into tertiles (first tertile ≈ ≤20 TPM; third tertile ≈ ≥60 TPM). Cases that fell between the two tertiles were labeled as ambiguous. In the combined cohort, no ambiguous cases remained after using GATA6 expression filtration. Samples in the lowest tertile were labeled basal-like, while the samples in the highest tertile were labeled classical. This refinement reflects clinical practice in which classical-like tumors with preserved differentiation (high GATA6) are considered biologically distinct from poorly differentiated, basal-like tumors.

After ssGSEA scoring, z-scoring, and GATA6 refinement, PANCAN (n = 614) contributed 176 high-confidence cases, out of which 99 were high-confidence basal-like, while 77 were high-confidence classical. On the other hand, TCGA (n = 183) contributed 62 high-confidence cases, out of which 15 were high-confidence basal-like, while 47 were high-confidence classical. These 238 high-confidence samples (176 PANCAN, 62 TCGA) served as the ground-truth for supervised model training and external testing. The resulting subtype proportions and classical/basal ratios were consistent with published PDAC transcriptomic datasets and maintain biological fidelity for model development.

### Overview of PanSubNet architecture

We propose PanSubNet, a deep learning framework that implements the language of histopathology, as proposed in our recent study [23], by integrating information across multiple scales and explicitly modeling cellular and intercellular characteristics within their biological context. The architecture is composed of three core components that mirror natural language processing (NLP) architectures: (1) multi-scale feature extraction to capture both cellular “words” and tissue patch “sentences”; (2) cell-to-patch mapping and fusion to create contextual embeddings; and (3) multi-dimensional attention mechanisms to learn complex semantic relationships between tissue regions.

Our strategy implements a dual-scale approach, where cells at 40× magnification serve as fundamental “words” encoding fine-grained morphological and phenotypic information, and tissue patches at 20× magnification represent “sentences” providing architectural context and spatial organization. Each WSI was tiled into non-overlapping 256 x 256-pixel patches at 20× magnification, and for each patch a feature vector was extracted using the pre-trained foundation model UNI2-h [39], capturing broad morphological and tissue-architectural patterns. Concurrently, cell nuclei were segmented and classified from WSIs at 40× magnification using CellViT++, specifically the SAM-based model [40–42]. This model yields embeddings for each individual cell, encoding fine-grained morphological and phenotypic details across five major cell categories, along with centroid coordinates for each cell.

To implement contextual embedding principles, we established a spatial correspondence between cells and patches by mapping each cell to the patch containing its centroid. Within each patch, we aggregated variable-length cell embeddings using a spatially biased self-attention mechanism with a learnable CLS token [43]. To incorporate spatial information, the physical Euclidean distance between cell centroids was subtracted from the raw attention scores. This spatial bias encodes biological function, where cellular proximity defines semantic relationships and functional neighborhoods. The CLS token was extracted, containing aggregated information from all cells within the patch with greater influence from spatially proximal cells.

To integrate cellular information with tissue architecture, we employed an outer product operation between the patch embedding vector and the aggregated cell embedding vector (CLS token) for each patch. This resulted in a matrix that captures all pairwise multiplicative interactions between patch and cell features. This representation was flattened and projected back to 768 dimensions using a learnable projection matrix to obtain the final fused representation. This fusion mechanism enables rich cross-scale interactions that capture how cellular characteristics interact with tissue architecture.

The fused embeddings from a WSI form a “bag” of k instances, which were aggregated using 2D AttMIL for slide-level prediction, as proposed in our recent study [23].

### Model training and evaluation

PanSubNet was trained exclusively on the 176 high-confidence PANCAN cases using five-fold cross-validation. In each fold, the model was trained to classify basal-like versus classical subtypes using binary cross-entropy loss, optimized with AdamW optimizer (learning rate 5e-05, weight decay 1e-05). Models were trained for up to 100 epochs with early stopping based on validation AUC. The final reported metrics represent the mean and standard deviation across all five folds.

For external validation, the model having best validation performance among five-fold PANCAN validation sets, was evaluated on the 62 high-confidence TCGA cases without any fine-tuning or retraining. This zero-shot evaluation assessed true cross-institutional generalizability. Performance metrics included area under the receiver operating characteristic curve (AUC), accuracy, balanced accuracy, sensitivity (recall for basal-like), and specificity (recall for classical).

### Baseline comparison

We compared PanSubNet against AttMIL [44], a standard architecture using UNI2-h patch embeddings without cellular information. AttMIL aggregates patch-level features through a single attention layer followed by a classification head. This baseline was trained and evaluated using identical data splits and preprocessing to enable direct comparison.

### Survival analysis

Kaplan–Meier survival analysis was performed in the PANCAN cohort to evaluate associations between molecular subtypes and OS. Analyses were conducted under multiple stratification settings, including patients who developed metastatic disease and the full cohort comprising both metastatic and resected patients. Molecular subtype assignments were obtained using either RNA-seq–derived molecular labels or PanSubNet model–predicted labels, enabling direct side-by-side comparison of labeling strategies. OS was defined as the time interval from the date of initial diagnosis to the date of death. Patients with documented death events were included in the analysis, while patients who were alive but lost to follow-up were excluded. Survival times were measured in months. Survival curves were estimated using the Kaplan–Meier method, and differences between groups were assessed using the log-rank test. Median overall survival values and corresponding 95% confidence intervals were computed for each group and reported in the figures. To evaluate the impact of subtype confidence, survival analyses were performed both in high-confidence molecular subtype cases and in the full PANCAN cohort including high- and low-confidence cases.

### Gene ontology and pathway enrichment analysis

Gene Ontology (GO) Biological Process and KEGG pathway enrichment analyses were performed on the top 25 most discriminative genes for each subtype (classical and basal-like) using the clusterProfiler R package. Enrichment p-values were calculated using hypergeometric tests with Benjamini–Hochberg correction for multiple testing. Significantly enriched terms (adjusted p < 0.05) were visualized as dot plots with −log10(p-value) on the x-axis. Gene-to-term networks were constructed using the enrichplot package to visualize the relationships between genes and their associated GO terms or KEGG pathways. These analyses can be found in Supplementary Materials.

### DNA damage repair gene expression analysis

Expression of key DDR genes (BRCA1, BRCA2, PALB2, RAD51, ATM, CHEK1) accounting for mutations, was extracted from the TPM matrices for all samples. A composite DDR score was calculated by z-scoring each gene across all samples and summing the z-scores. DDR gene expression was compared between subtypes using violin plots and heatmaps. Correlation analyses assessed the relationship between DDR score and subtype confidence (ΔZ) and prediction ambiguity. These analyses can be found in the Supplementary Materials.

## Results

### Construction of high-confidence molecular labels across PANCAN and TCGA cohorts

We first derived transcriptomic ground-truth labels for molecular subtyping using Moffitt 50-gene basal-like and classical expression using techniques derived from previous literature [45, 46]. Single-sample gene-set enrichment analysis (ssGSEA) was used to compute subtype enrichment, and samples with |z-score| > 1 were designated as high-confidence basal-like or classical subtypes [47]. Intermediate cases were retained for downstream exploration but excluded from supervised model training.

Across the Pancreatic Cancer Action Network (PANCAN) cohort (n = 846), molecular data were available for 614 cases, including 176 high-confidence labels (99 basal-like and 77 classical) [48]. The 176 high-confidence samples were stratified based on clinical disease status, comprising 66 metastatic cases and 110 surgically resected cases.. Within The Cancer Genome Atlas (TCGA) cohort (n = 209), molecular data were available for 183 cases, with 62 high-confidence classifications (15 basal-like and 47 classical) (see **Supplementary Figure S1**). These high-confidence subsets formed the basis for model training, internal validation, and external testing. The distribution of high-confidence subtypes was consistent with previously reported proportions for PDAC transcriptomic classes and reflects the predominance of the classical lineage in bulk RNA sequencing datasets [49].

### PanSubNet accurately predicts basal-like and classical subtypes in internal validation on PANCAN

PanSubNet was trained exclusively on the high-confidence PANCAN cases and evaluated using a five-fold cross-validation framework. The model demonstrated strong discriminatory ability, achieving a mean AUC of 88.514 ± 5.344 and an accuracy of 84.912 ± 10.050 across folds (see **Figure 2, Supplementary Table S1**). Balanced accuracy (85.219 ± 10.227) indicated that performance was stable across both subtypes, with no substantial bias despite the modest class imbalance. Sensitivity and specificity were similarly well-aligned (84.602 ± 13.218 and 85.837 ± 8.969, respectively), suggesting that PanSubNet reliably distinguishes basal-like from classical tumors even within a biologically heterogeneous cohort.

**Figure 2.**
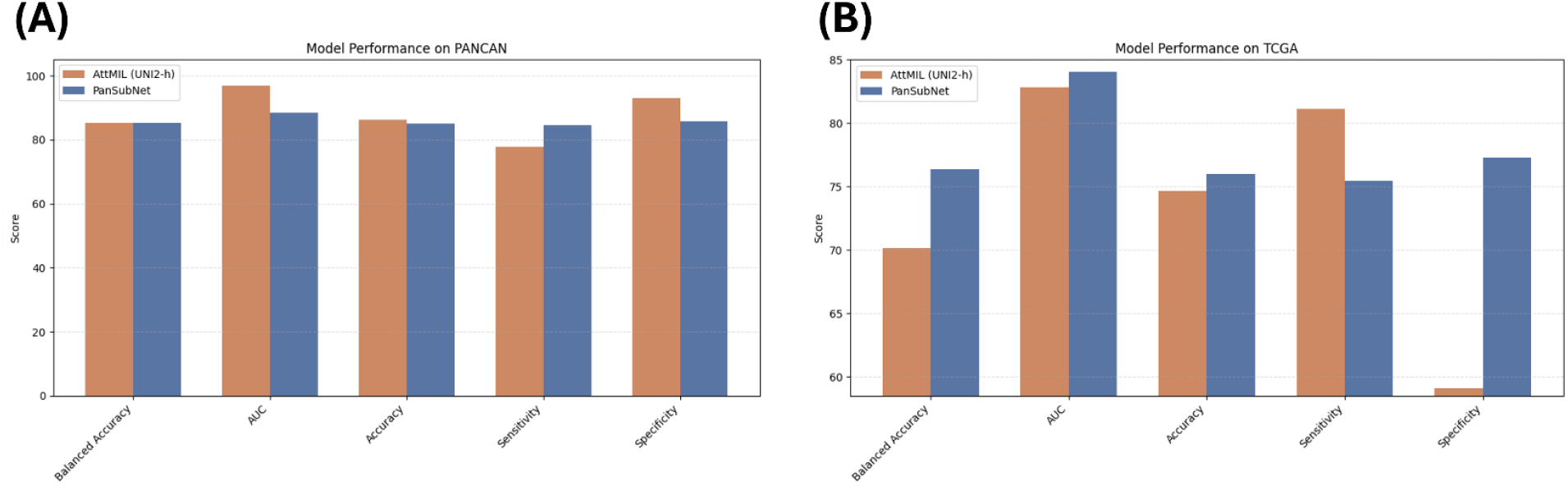
Performance comparison of PanSubNet and AttMIL across internal and external cohorts. Comparison of PanSubNet and AttMIL (UNI2-h backbone) performance on the PANCAN and TCGA cohorts. Metrics shown include area under the receiver operating characteristic curve (AUC), accuracy, balanced accuracy, sensitivity, and specificity.

To assess PanSubNet performance in a more realistic and transcriptomically ambiguous setting, we conducted an exploratory analysis in which the trained model, learned exclusively from high-confidence PANCAN cases, was applied at inference time to the entire PANCAN cohort, including samples with low-confidence subtype scores. As anticipated, the inclusion of biologically ambiguous tumors resulted in a measurable reduction in predictive performance. Across five-fold cross-validation, PanSubNet achieved an AUC of 71.332 ± 6.113, accuracy of 68.106 ± 4.683, balanced accuracy of 68.045 ± 4.807, sensitivity of 70.078 ± 5.927, and specificity of 66.012 ± 7.934 (see **Supplementary Table S1**).

Across 189 samples, the model achieved an overall concordance (accuracy) of 0.847, with 29 misclassifications. Error rates were comparable between subtypes, with 14/99 errors in basal tumors (14.1%) and 15/90 errors in classical tumors (16.7%), indicating no strong class-specific bias. Correct predictions exhibited significantly higher decision margins than incorrect predictions (median margin 0.462 vs. 0.384; Mann–Whitney U test, p = 0.014), suggesting that misclassifications are associated with reduced model confidence. Notably, 13 of 29 errors (44.8%) occurred with high predicted confidence (p ≤ 0.10 or ≥ 0.90), consistent with a subset of confidently misclassified cases rather than uniformly ambiguous predictions. Concordance of RNA sequencing and PanSubNet is discussed in **Supplementary Table S3**.

### PanSubNet demonstrates strong generalizability in external validation on TCGA

To assess cross-cohort generalizability, PanSubNet trained exclusively on PANCAN WSIs was evaluated on the high-confidence TCGA PDAC cohort without any fine-tuning. Despite differences in institutional origin, sequencing platforms, slide preparation, and staining variability, the model maintained high performance, achieving an AUC of 84.048 and accuracy of 76.000 (see **Figure 2, Supplementary Table S1**). Balanced accuracy (76.372) demonstrated that classification remained stable across subtypes, and sensitivity and specificity (75.472 and 77.273, respectively) confirmed robust external performance.

### Benchmarking against baseline reveals superior cross-cohort stability of PanSubNet

We compared PanSubNet against the attention-based multiple instance learning model AttMIL with UNI2-h, a widely used WSI encoder [39, 44]. On internal evaluation using PANCAN, AttMIL achieved a similar balanced accuracy (85.336 ± 4.718) and higher AUC (96.812 ± 1.026), but at the expense of a substantial imbalance between sensitivity and specificity. This imbalance reflects a bias toward predicting the classical subtype, resulting in under-calling basal-like cases. In external validation on TCGA, AttMIL exhibited a marked reduction in performance, with AUC decreasing to 82.847, accuracy to 74.667, and specificity to 59.091. In contrast, PanSubNet preserved high discriminative performance on TCGA, with stable sensitivity and specificity across cohorts (see **Figure 2, Supplementary Table S2)**.

### Molecular subtypes stratify patients by overall survival in the PANCAN cohort

We evaluated the association between molecular subtypes and overall survival (OS) in the PANCAN cohort using Kaplan–Meier analysis under multiple stratification settings (see Figure 3). Analyses were performed separately for patients who developed metastatic disease and for the full cohort, including both metastatic and resected patients. Subtype assignments were derived from both RNA-seq–based molecular labels and PanSubNet model predictions to enable side-by-side comparison.

**Figure 3.**
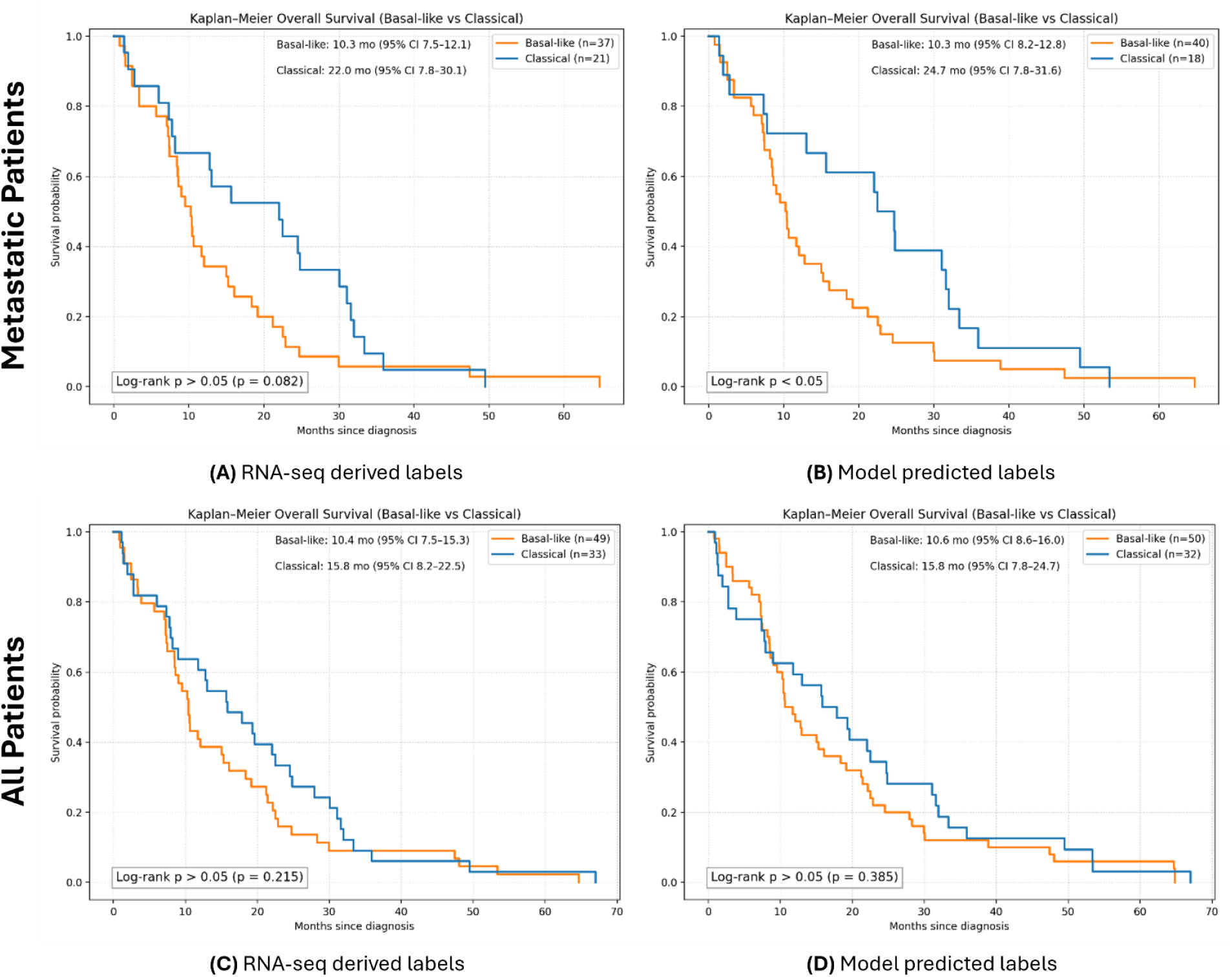
Overall survival stratified by molecular subtype in the PANCAN cohort (high-confidence cases). Kaplan–Meier curves showing overall survival (OS) stratified by Basal-like and Classical subtypes across different patient subsets and labeling strategies. **(A)** OS among patients who developed metastatic disease, stratified using RNA-seq–derived molecular subtype labels. **(B)** OS among the same metastatic patients, stratified using PanSubNet-predicted molecular subtypes. **(C)** OS in all high confidence cases (metastatic and resected), stratified using RNA-seq–derived molecular subtype labels. **(D)** OS in all high confidence cases, stratified using PanSubNet-predicted molecular subtypes. Median survival times with 95% confidence intervals are reported within each panel. Log-rank test results are indicated on the plots.

Among metastatic patients, OS stratification was first evaluated using high-confidence RNA-seq–derived molecular subtype labels. In this high-confidence subset, Kaplan–Meier analysis showed separation between classical and basal-like tumors; the difference had a trend towards statistical significance (p=0.08, see **Figure 3a**). When the same metastatic subset was stratified using PanSubNet-predicted subtypes, overall survival differed significantly between groups (see **Figure 3b**). Notably, several cases labeled as classical by RNA-seq were predicted as basal-like by the model, and these misclassified cases experienced early death events. Consequently, the median OS of the classical group differed between labeling strategies, with a median OS of 22.0 months using RNA-seq–derived labels compared to 24.7 months using model-predicted labels. This preservation of survival stratification using histology-only inference indicates that PanSubNet captures clinically relevant tumor biology within routine WSIs.

We next assessed OS in all high-confidence cases, including both metastatic and resected patients. In this setting, RNA-seq–derived molecular subtypes demonstrated modest separation in OS that did not consistently reach statistical significance (see **Figure 3c**). Stratification of the same cohort using PanSubNet-predicted subtypes likewise showed limited separation between groups, with no statistically significant differences observed (see **Figure 3d**).

To examine the impact of subtype confidence on survival associations, we additionally repeated OS analyses in the full PANCAN cohort, including both high- and low-confidence cases (see **Supplementary Figure S2**). Across these analyses, inclusion of low-confidence cases was associated with reduced separation between survival curves compared to analyses restricted to high-confidence molecular labels. The confidence-dependent improvement in the prognostic signal suggests that internal estimates meaningfully reflect biological fidelity rather than model noise.

In summary, survival differences between RNA-sequencing–defined subtypes were more apparent in the metastatic subgroup than in the full cohort, which includes resected early-stage cases, highlighting the confounding influence of disease stage and surgical resection on overall survival. Notably, PanSubNet preserved robust prognostic stratification in metastatic patients and demonstrated stronger survival separation than RNA-seq–based subtype labels in this setting.

## Discussion

Our findings demonstrate that the basal-like and classical subtypes of PDAC, originally defined through transcriptomic profiling, manifest as distinct and morphologically learnable histopathological patterns. This observation is consistent with fundamental principles of tumor biology: transcriptional programs shape cellular phenotypes, and these phenotypes leave measurable morphological signatures within tissue architecture that can be captured in routine H&E-stained sections [50]. The ability of PanSubNet to infer RNA-seq–defined molecular subtypes directly from histology supports the premise that transcriptional identity is encoded in tissue organization, cellular morphology, and microenvironmental context. This, to our knowledge, represents the first WSI-based PDAC subtyping according to the Moffitt classification system. Prior studies were based on PurIST classifier, which demonstrates a incomplete concordance with the Moffitt system [15, 32, 35, 51]. A foundational step of this study was establishing and validating the molecular ground truth used to train PanSubNet. Since PanSubNet is explicitly optimized to predict RNA-sequencing–derived basal-like versus classical subtype labels, all transcriptomic analyses were intentionally performed using RNA-seq–derived subtype assignments rather than model-predicted labels. This design avoids circular inference: using PanSubNet predictions to characterize transcriptomic programs would confound biological interpretation, as any observed differences would primarily reflect model optimization rather than independent molecular structure. Accordingly, RNA-seq analyses in this work serve exclusively to confirm that the supervised target labels correspond to coherent biological programs and clinically meaningful phenotypes, rather than to introduce a parallel molecular discovery effort. Detailed transcriptomic characterizations, including lineage markers, pathway enrichment, and subtype ambiguity analyses, are therefore presented in the **Supplementary Methods 1-6**, while the primary Results section focuses on histology-based prediction performance and clinical associations.

Within these RNA-seq–based validation analyses, GATA6 emerged as a robust lineage-associated marker, exhibiting elevated expression in classical tumors and reduced or absent expression in basal-like tumors, including a subset of samples with undetectable expression (**Supplementary Methods 3**). This distribution is consistent with prior literature and reinforces GATA6’s role as a marker of pancreatic epithelial differentiation. Importantly, GATA6 is not proposed here as an alternative subtyping framework, but rather as a biological anchor that corroborates the transcriptional identity of the classical subtype and helps contextualize intermediate or ambiguous cases [16, 33]. Together with pathway-level analyses, these findings confirm that the RNA-seq– derived subtype labels used for model supervision represent stable, biologically grounded endpoints.

Ambiguity analyses further revealed that PDAC molecular subtypes exist along a continuous transcriptional spectrum, with basal-like and classical tumors occupying relatively stable endpoints and intermediate cases forming a transitional zone. This biological continuum presents an inherent challenge for supervised classification, as no sharp boundary separates subtypes. By training exclusively on high-confidence cases, where transcriptional identity is unambiguous, PanSubNet learns the clearest morphological correlates of the subtype extremes. The model’s attention mechanisms then enable interpolation across this spectrum, capturing patterns consistent with partial differentiation or incomplete dedifferentiation. The strong external validation performance across institutions, staining protocols, and specimen types suggests that these learned patterns generalize beyond cohort-specific artifacts (see **Figures 4 and 5**).

**Figure 4.**
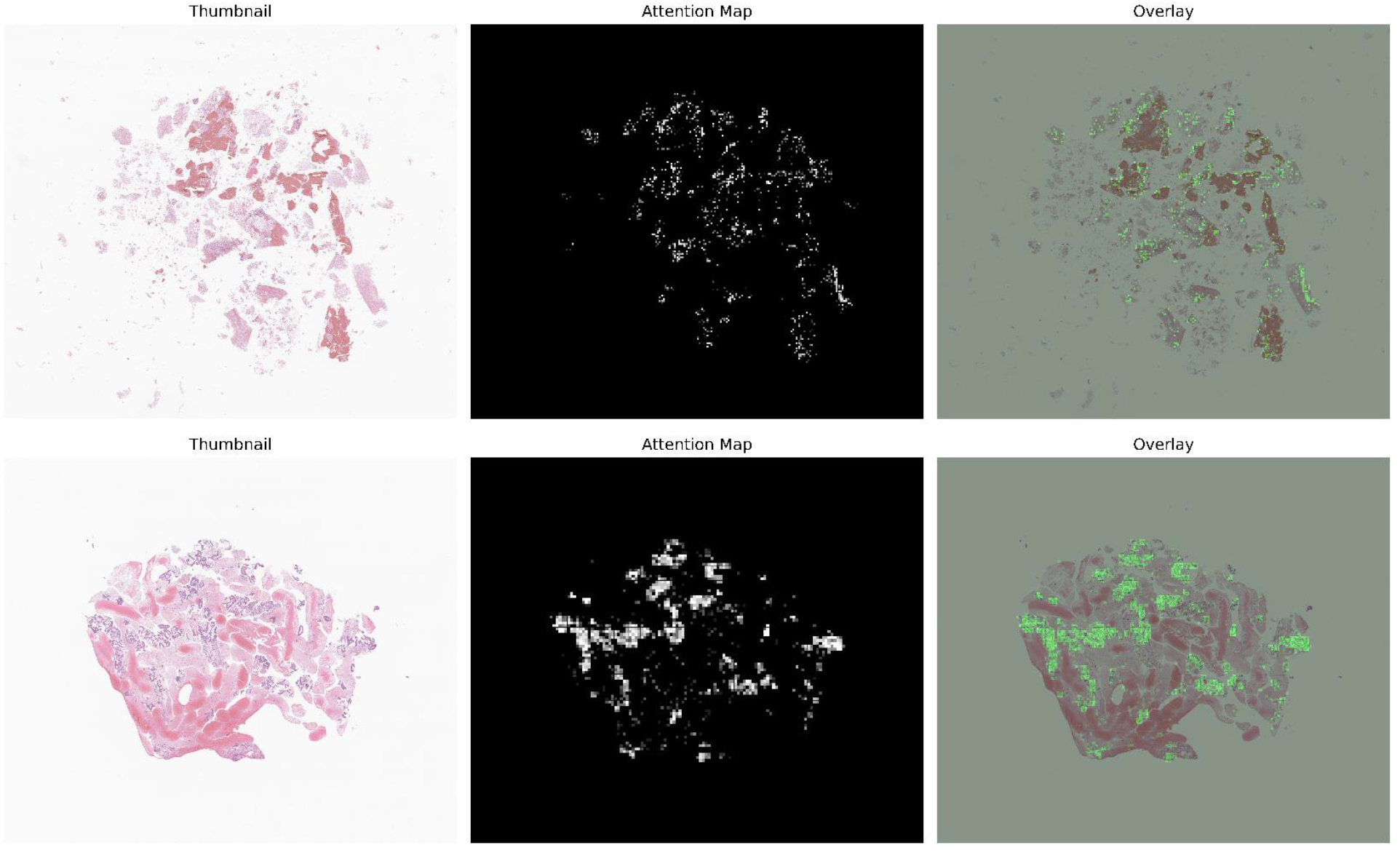
PanSubNet attention maps highlight discriminative regions in classical subtype patients. Attention maps from PanSubNet for fine needle biopsy specimens, from two randomly sampled, correctly predicted classical-subtype patients. The left panel shows a thumbnail of the original whole-slide image. The middle panel displays the attention mask, with values ranging from 0 (black) to 255 (white), where white indicates the highest attention. The right panel shows the attention map overlaid on the original slide.

**Figure 5.**
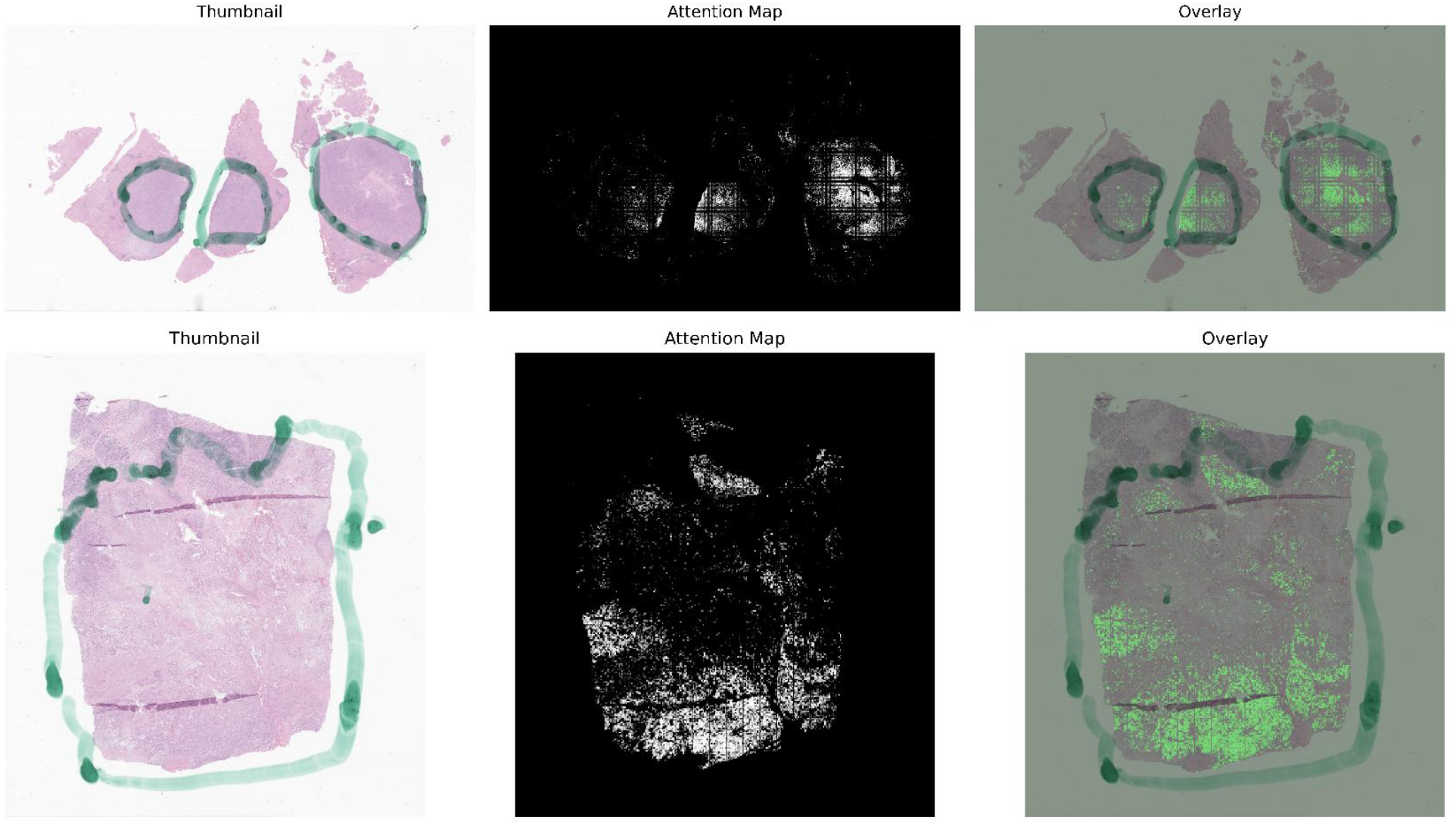
PanSubNet attention maps identify basal-like–associated regions. Attention maps from PanSubNet for surgical resection specimens, from two randomly sampled, correctly predicted basal-like-subtype patients. The left panel shows a thumbnail of the original whole-slide image. The middle panel displays the attention mask, with values ranging from 0 (black) to 255 (white), where white indicates the highest attention. The right panel shows the attention map overlaid on the original slide. The region marked in green, visible in some slides, was not used during model training and appeared randomly across patients regardless of subtype; in this case, the two basal-like examples happened to contain this mark by chance.

Consistent with prior studies, classically subtyped tumors by RNA-sequencing exhibited more favorable survival than basal-like tumors, with a trend towards significance in metastatic patients (p=0.08) [52]. Our analyses reveal that subtype differences are most pronounced in metastatic cases, where tumor biology exerts an influence outcome. PanSubNet subtype assignments maintained, and even improved survival stratification compared to RNA-seq labels. Some tumors labeled classical by RNA-seq but basal-like by PanSubNet, which had early deaths, suggest histomorphology at the whole-slide level reflects aggressive biology not fully captured by bulk transcriptomics. Including resected patients reduced survival differences across subtypes for both methods, highlighting the influence of surgery and disease stage on long-term survival outcomes. Transcriptomic analyses linking subtype status with DNA damage repair (DDR) gene expression provide a plausible biological rationale for these clinical differences and suggest that integrating subtype information with DDR-related biomarkers may ultimately support more refined therapeutic stratification (**Supplementary Methods 5**). However, such integrative approaches remain exploratory and were not evaluated directly in this study.

PanSubNet represents a step toward democratizing molecular stratification in PDAC. Unlike RNA sequencing, which requires specialized infrastructure, sufficient tissue yield, and prolonged turnaround, PanSubNet operates directly on H&E slides, the standard output of diagnostic pathology workflows worldwide. WSIs can be scanned, analyzed, and classified within hours, offering subtype information on a clinically relevant timescale. This rapid turnaround is particularly important in PDAC, where disease progression often necessitates urgent treatment decisions.

The model’s robust external performance on TCGA (AUC 0.84) demonstrates resilience to inter-institutional variability, an essential prerequisite for clinical deployment. Notably, PanSubNet maintained balanced sensitivity and specificity across cohorts, avoiding the classical-subtype bias observed in the AttMIL baseline. This suggests that the multi-scale, cell-to-patch fusion architecture more effectively captures biologically meaningful features than patch-level aggregation alone. Misclassification rates were similar for basal-like and classical tumors, with errors mainly in ambiguous cases, not systematic bias. Some discordant cases likely show true histologic– transcriptomic divergence, possibly due to heterogeneity, sampling, or lineage plasticity. These findings support PanSubNet as a robust, biologically grounded WSI-based subtyping approach.

Several limitations warrant consideration. The ground-truth labels were derived using the Moffitt 50-gene signature with ssGSEA scoring. While well-validated, alternative PDAC subtyping frameworks such as PurIST exist, and the optimal classification schema remains debated [53–55]. Training PanSubNet using alternative label definitions may clarify whether the model learns generalizable morphological correlates of tumor biology or schema-specific patterns [56].Although PanSubNet was developed and evaluated in a cohort exceeding 1000 patients, PDAC is a biologically and histologically heterogeneous disease; therefore, further validation in larger, more diverse, multi-institutional datasets will be important for assessing generalizability.

We acknowledge that molecular subtyping alone has limited immediate impact on treatment selection in the current PDAC therapeutic landscape, where first-line options remain restricted. Nevertheless, subtype assignment provides meaningful prognostic context that may support anticipatory clinical planning. For basal-like tumors, this includes early recognition of aggressive biology and prioritization of clinical trial enrollment; for classical tumors, it may support toxicity-sparing strategies and sustained therapy continuation. In its current iteration, PanSubNet does not currently integrate clinical variables or immunohistochemical markers. Hybrid models combining histology-based inference with targeted IHC or clinical features may improve interpretability and risk stratification but are beyond the scope of this work. Future work will focus on prospective validation, integration with orthogonal biomarkers, and extension of histology-based molecular inference to additional tumor types where transcriptional subtyping informs therapy response [57]PanSubNet is intended to complement current clinical and pathological evaluations, and is not designed to provide treatment recommendations or function as a clinical decision-support tool.

In summary, our PanSubNet classifies PDACs into clinically useful subtypes directly from the routine H&E slides with biological and clinical fidelity. By lowering barriers to molecular stratification, it enables scalable integration of tumor biology into translational research and clinical workflows. clinical contexts where transcriptomic profiling is impractical, including small biopsies with insufficient RNA yield, community hospitals without access to sequencing platforms, rapid triage in metastatic disease, retrospective stratification of archival specimens for clinical trials, and intraoperative or cytology specimens.

## Code and data availability

The underlying code for this study is available in AI4Path PanSubNet repository and can be accessed via this link https://github.com/AI4Path-Lab/PanSubNet. WSIs and RNA-seq data for TCGA-PAAD are publicly available through the Genomic Data Commons (https://portal.gdc.cancer.gov/). PANCAN data access is subject to institutional data use agreements. Processed data and model weights will be made available upon reasonable request to the corresponding author.

## Author Contributions

A.R.A. and A.L. contributed equally to this work. A.R.A. designed and implemented the PanSubNet architecture, performed computational analyses, and wrote the manuscript. A.L. performed RNA sequencing preprocessing, molecular characterization, gene ontology analyses, and contributed to manuscript writing. A.E. provided pathology expertise, reviewed the attention maps of the model, and contributed to manuscript editing and revision. A.P. and E.H. provided oversight and resources for the study and contributed to manuscript editing and revision. W.C. supervised the clinical side of the study, participated in model training to ensure clinical relevance, and contributed to manuscript editing and revision. A.M. contributed to study conception, study design, and analytical strategy, provided expert guidance on pancreatic cancer biology, molecular subtyping, and clinical relevance, and contributed to manuscript writing, editing, and revision. M.K.K.N. conceptualized, designed, and validated the study, supervised the research, provided funding, and edited and revised the manuscript.

## Competing Interests

The authors declare no competing interests.

## Acknowledgments

We thank the patients who contributed samples to the PANCAN and TCGA cohorts. We acknowledge the Genomic Data Commons for providing public access to TCGA data. We also gratefully acknowledge the Ohio Supercomputer Center for providing high-performance computing resources as part of its contract with The Ohio State University College of Medicine. We also thank the Department of Pathology and the Comprehensive Cancer Center at The Ohio State University for their support.

## Funding

The project described was supported in part by R01 CA276301 (PIs: Niazi and Chen) from the National Cancer Institute, Pelotonia under IRP CC13702 (PIs: Niazi, Vilgelm, and Roy), The Ohio State University Department of Pathology and Comprehensive Cancer Center. The content is solely the responsibility of the authors and does not necessarily represent the official views of the National Cancer Institute or National Institutes of Health or The Ohio State University.

## Ethics Approval and Consent to Participate

This study involved secondary analysis of retrospective, fully de-identified clinical, molecular, and histopathology data obtained from existing institutional and public repositories. In accordance with applicable regulations, the use of de-identified data does not constitute human subjects’ research. Therefore, Institutional Review Board (IRB) approval was not required, and informed consent to participate was waived.

## Notes

### Competing Interest Statement

The authors have declared no competing interest.

