## Supplementary material for "Inferring Clinically Relevant Molecular Subtypes of Pancreatic Cancer from Routine Histopathology Using Deep Learning": Supplemetal Table S1

**Supplementary Materials**

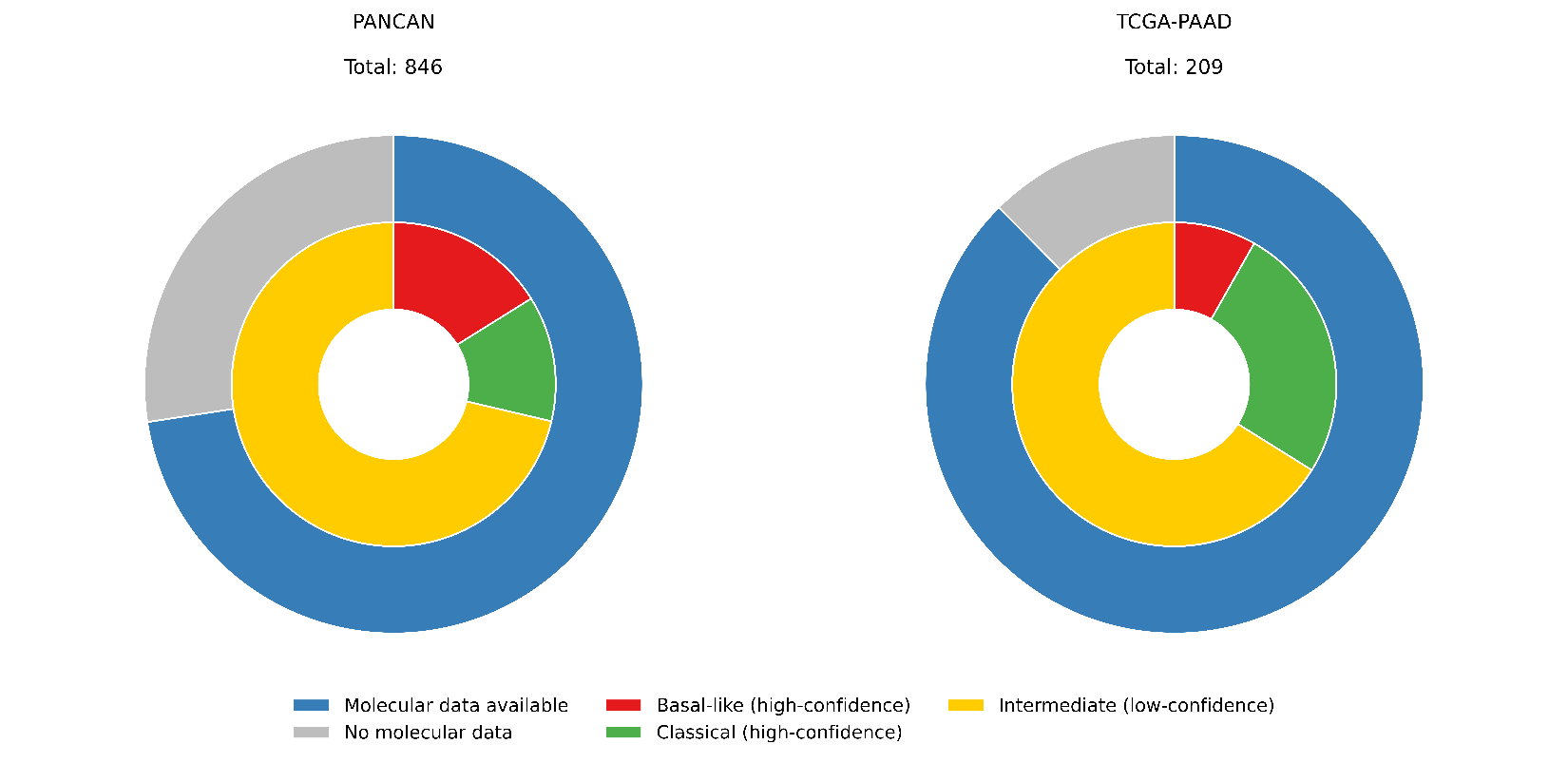

***Figure S1:*** *Nested donut charts illustrating cohort composition and molecular subtype distribution. The outer ring represents the proportion of patients with available molecular data (blue) versus those without (gray) for each cohort (PANCAN and TCGA-PAAD). The inner ring shows the breakdown of molecular cases into high-confidence subtypes: basal-like (red) and classical (green) and intermediate/low-confidence cases (yellow).*

***Table S1. Performance of PanSubNet for molecular subtype prediction across cohorts.***

| **Dataset** | **AUC** | **Accuracy** | **Bal. Acc.** | **Sensitivity** | **Specificity** |
| --- | --- | --- | --- | --- | --- |
| **PANCAN (high confidence)** | 88.514 ± 5.344 | 84.912 ± 10.050 | 85.219 ± 10.227 | 84.602 ± 13.218 | 85.837 ± 8.969 |
| **PANCAN (entire cohort)** | 71.332 ± 6.113 | 68.106 ± 4.683 | 68.045 ± 4.807 | 70.078 ± 5.927 | 66.012 ± 7.934 |
| **TCGA** | 84.048 | 76.000 | 76.372 | 75.472 | 77.273 |

*The table summarizes classification performance of PanSubNet on the PANCAN cohort using high-confidence RNA-seq–derived labels, the entire PANCAN cohort (including lower-confidence cases), and the independent TCGA cohort. Reported metrics include area under the ROC curve (AUC), accuracy, balanced accuracy (Bal. Acc.), sensitivity, and specificity. Values for PANCAN are shown as mean ± standard deviation across cross-validation folds, while TCGA results reflect single-run external validation.*

***Table S2. Performance of AttMIL (UNI2-h backbone) for molecular subtype prediction.***

| **Dataset** | **AUC** | **Accuracy** | **Bal. Acc.** | **Sensitivity** | **Specificity** |
| --- | --- | --- | --- | --- | --- |
| **PANCAN (high confidence)** | 96.812 ± 1.026 | 86.131 ± 3.571 | 85.336 ± 4.718 | 77.678 ± 12.443 | 92.995 ± 6.783 |
| **TCGA** | 82.847 | 74.667 | 70.111 | 81.132 | 59.091 |

*This table reports the performance of the attention-based multiple instance learning (AttMIL) model with the UNI2-h feature extractor on the PANCAN high-confidence cohort and the external TCGA cohort. Metrics include AUC, accuracy, balanced accuracy, sensitivity, and specificity. PANCAN results are reported as mean ± standard deviation across cross-validation folds, whereas TCGA results correspond to external validation performance.*

***Table S3. Concordance of RNA sequencing with PanSubNet in TCGA database***

| **Metric** | **Result** | **Interpretation** |
| --- | --- | --- |
| Total samples | 189 | Evaluation cohort |
| Overall concordance (accuracy) | 0.847 | High agreement between PanSubNet predictions and reference labels |
| Total misclassifications | 29 | Errors across both subtypes  Comparable error rate  No subtype-specific bias |
| Basal-like errors | 14 / 99 (14.1%) |  |
| Classical errors | 15 / 90 (16.7%) |  |
| Decision margin (correct predictions, median) | 0.462 | Higher model confidence |
| Decision margin (incorrect predictions, median) | 0.384 | Lower confidence |
| Statistical comparison of margins | *p* = 0.014 (Mann–Whitney U) | Errors associated with reduced confidence |
| High-confidence errors | 13 / 29 (44.8%) | Subset of confidently misclassified cases |
| High-confidence definition | ≤ 0.10 or ≥ 0.90 | Confidence threshold |

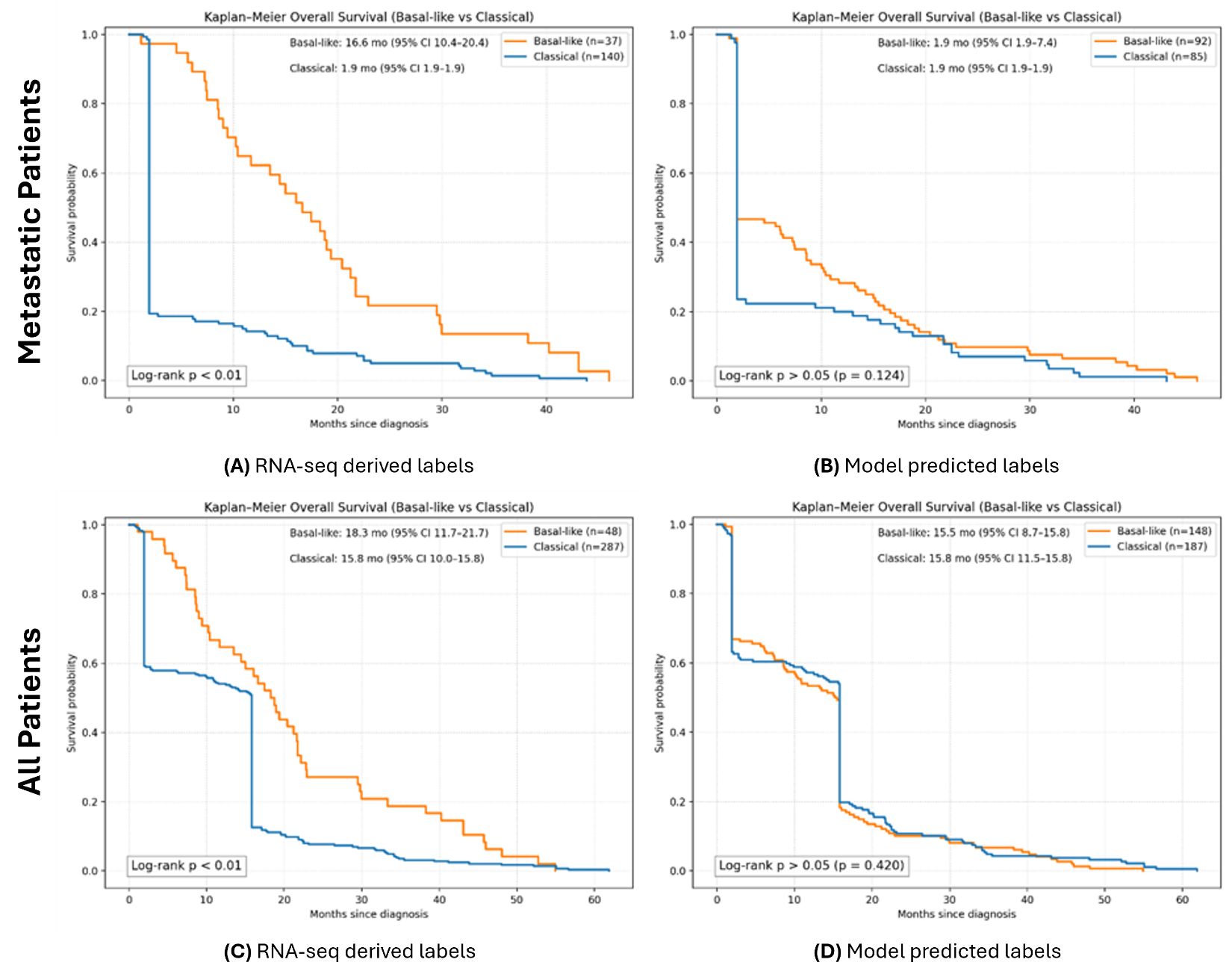

***Supplementary Figure S2. Overall survival stratified by molecular subtype in the PANCAN cohort including all cases (high- and low-confidence).*** *Kaplan–Meier curves showing overall survival (OS) stratified by Basal-like and Classical subtypes including both high- and low-confidence cases.* ***(A)*** *Metastatic patients stratified by RNA-seq–derived labels.* ***(B)*** *Metastatic patients stratified by PanSubNet-predicted labels.* ***(C)*** *Full cohort stratified by RNA-seq–derived labels.* ***(D)*** *Full cohort stratified by PanSubNet-predicted labels. Median survival times with 95% confidence intervals and log-rank test results are shown for each comparison.*

**Supplementary Methods 1: Establishing the validity of ground truth labels through transcriptomic analyses**

All transcriptomic analyses referenced in this study were performed using RNA-sequencing–derived basal-like versus classical subtype labels, and this choice was made intentionally. These RNA-seq analyses are included solely to establish that the subtype labels PanSubNet is trained to predict correspond to coherent biological programs and clinically meaningful outcomes, rather than to introduce an independent or parallel molecular analysis. PanSubNet is an H&E-based model whose sole objective is to predict the RNA-seq–defined basal-like versus classical subtype. It does not propose, redefine, or generate an independent histology-derived molecular taxonomy. Instead, the model learns histologic patterns that best approximate an established and biologically validated transcriptomic ground truth.

Accordingly, using PanSubNet-derived subtype predictions for transcriptomic characterization would not be viable. The model’s predictions are explicitly optimized to reproduce RNA-seq–defined labels, and reusing those predictions to stratify gene expression would not constitute an independent biological validation. Within this framework, PanSubNet does not redefine subtype biology. Rather, it learns morphological correlates that approximate a previously established molecular phenotype. The transcriptomic analyses therefore serve as a biological anchor that justifies the prediction target itself, rather than as a discovery analysis intended to compete with the image-based model.

In this context, the image modality and the RNA-seq labels are not statistically independent, nor are they intended to be. What is independent, and what this study explicitly interrogates, is the set of histologic features learned by the model to recover the molecular subtype from routine H&E slides. The RNA-seq analyses are included to validate the biological and clinical relevance of the target label, not to supersede or parallel the image-based results. The rationale is to first establish that RNA-seq–defined basal-like and classical subtypes correspond to coherent differentiation programs, pathway activity, and prognostic behavior, and then to demonstrate that PanSubNet can accurately predict these subtypes directly from histology.

**Supplementary Methods 2: Molecular characterization reveals distinct transcriptomic programs anchored by differentiation state**

To validate the biological coherence of our ground-truth labels, we performed comprehensive molecular characterization of the classified subtypes. Principal component analysis using the top Moffitt genes demonstrated clear separation between subtypes (see Supplementary Figure S3c), with the primary component (PC1) capturing the classical-basal axis and accounting for the strongest variance. The secondary component (PC2) contributed 9% of variance, reflecting intra-subtype heterogeneity. Notably, samples clustered tightly with their respective subtype neighbors, indicating minimal transcriptomic ambiguity in high-confidence classifications.

Analysis of the most discriminative genes revealed distinct biological programs (see Supplementary Figure S3d). The classical subtype was characterized by high expression of trefoil factors (TFF1, TFF2, TFF3), which are critical for ductal differentiation and maintenance of the epithelial barrier [1]. The expression of REG4 indicated activation of PI3K/AKT and ERK/MAPK pathways, consistent with KRAS-driven oncogenesis in a differentiated cellular context [2]. Additional classical markers included SPINK1, reflecting trypsin inhibition and differentiated pancreatic function; TSPAN8 and LGALS4, involved in cellular organization and adhesion; and CEACAM5, AGR2, and CTSE, all indicators of mucin biology and functional epithelial organization [3-5].

In contrast, the basal-like cohort showed predominant expression of SCEL, an indicator of TP63 activity, and squamous differentiation with mesenchymal plasticity [6]. The expression of KLK8 and CST6 reflected loss of mucal organization and stromal infiltration, accompanied by matrix metalloproteinase activity [7, 8]. KRT6A and S100A2 signified squamous plasticity through extracellular matrix remodeling and wound repair processes [9].

Aggregate subtype program scores confirmed these patterns. Basal-like tumors showed significantly elevated basal program enrichment (see Supplementary Figure S3e), while classical tumors demonstrated higher classical program scores (see Supplementary Figure S3f). Importantly, the variance in basal marker expressions within classical tumors was greater than the variance in classical marker expressions within basal-like tumors.

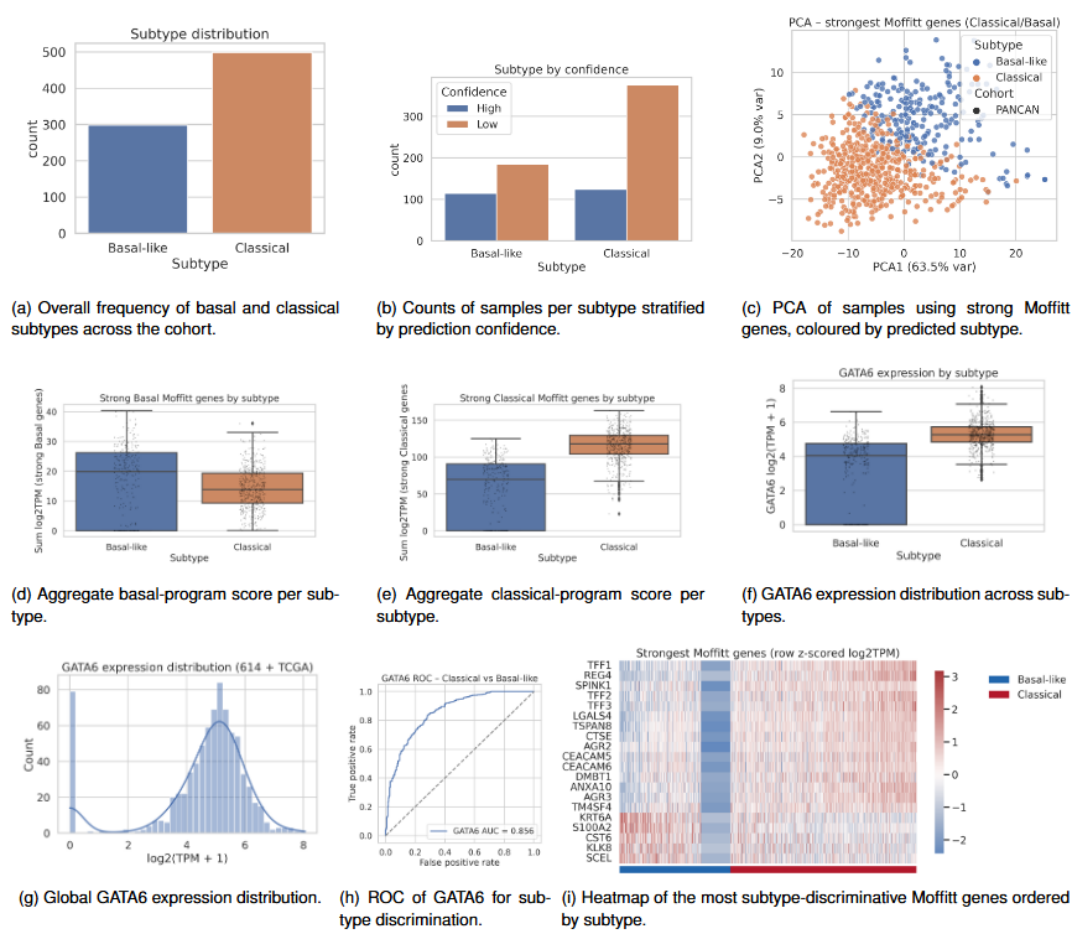

***Supplementary Figure S3:*** *Integrated molecular characterization of Basal-like and Classical subtypes using PANCAN + TCGA cohorts. This figure summarizes the distribution of predicted subtypes, subtype-specific gene expression patterns, and marker-based discrimination between Basal-like and Classical pancreatic cancer.* ***(A)*** *Subtype distribution: Bar plot showing the overall frequency of Basal-like and Classical cases across the combined cohort, with Classical tumors occurring more frequently.* ***(B)*** *Subtype by prediction confidence: Stratification of predictions into high- and low-confidence groups shows that high-confidence calls are more enriched within the Classical subtype, whereas Basal-like cases include a larger proportion of low-confidence classifications.* ***(C)*** *PCA using strongest Moffitt subtype-defining genes: Principal component analysis based on the most discriminative classical/basal genes from the Moffitt signature. Points are colored by predicted subtype and shaped by cohort (PANCAN vs TCGA). Classical and Basal-like tumors show clear separation primarily along PC1. .* ***(D)*** *Basal Moffitt gene expression: Boxplots of summed log2(TPM+1) expression for strong Basal-like genes demonstrate significantly higher expression in Basal-like tumors compared to Classical tumors.* ***(E)*** *Classical Moffitt gene expression: Boxplots of summed log2(TPM+1) expression for the strong Classical genes reveal strong enrichment in Classical tumors relative to Basal-like tumors.* ***(F)*** *GATA6 expression by subtype: Boxplot of GATA6 expression showing markedly elevated expression in Classical tumors, with lower and more heterogeneous expression in Basal-like tumors.* ***(G)*** *Overall GATA6 expression distribution: Histogram and kernel density estimate illustrating the unimodal but right-skewed distribution of GATA6 expression across the combined dataset.* ***(H)*** *ROC curve for GATA6 distinguishing Classical vs Basal-like: Receiver operating characteristic analysis demonstrating strong discriminative ability of GATA6 expression alone (AUC = 0.856) for separating Classical from Basal-like subtypes.* ***(I)*** *Heatmap of strongest Moffitt genes: Z-scored expression heatmap of key subtype driver genes highlights coherent Classical-high and Basal-high gene modules.*

**Supplementary Methods 3: GATA6 expression provides robust discrimination of molecular subtypes**

GATA6, a key transcription factor regulating pancreatic differentiation, showed markedly different expression patterns across subtypes (see Supplementary Figure S3g). GATA6 was used solely as a principled mechanism to clarify a small subset of transcriptionally ambiguous samples when enrichment-based Moffitt gene scores alone were insufficient to confidently assign classical or basal-like subtype. Importantly, GATA6 is not used here as a primary subtype classifier, nor is it proposed as a standalone alternative to established subtyping frameworks. Its role is limited to increasing subtype confidence within a narrowly defined ambiguous group, a use that is consistent with prior literature. In high-confidence TCGA samples, GATA6 expression was significantly higher in classical tumors than basal tumors (classical mean 4.94 vs basal mean 4.03; Mann–Whitney U p = 2.43 × 10⁻⁸, FDR-adjusted p = 1.94 × 10⁻⁷), consistent with its established role as a classical lineage marker. Classical tumors exhibited higher median GATA6 expression with lower variance, while basal-like tumors showed substantially reduced expression with greater spread. Notably, 80 samples across both cohorts showed no detectable GATA6 expression, predominantly within the basal-like group. Despite this, the majority of samples displayed moderate to high GATA6 levels.

The global distribution of GATA6 expression (see Supplementary Figure S3h) revealed a bimodal pattern consistent with its role as a lineage-defining factor. Remarkably, GATA6 expression alone achieved an AUC of 0.86 for discriminating classical from basal-like subtypes (see Supplementary Figure S3i), validating its utility as a single-gene classifier for ambiguous or intermediate-confidence cases (p <<.001, DeLong’s Test). Across 178 patients (93 events; basal n = 34, classical n = 144), GATA6 expression was not prognostic, with no significant difference in overall survival between high and low GATA6 groups (log-rank p = 0.971). In contrast, basal versus classical subtype status remained significantly associated with survival (log-rank p = 0.0145).This strong discriminative power supports the biological rationale for using GATA6 to refine intermediate classifications, as GATA6 activity is heavily correlated with classical phenotypes [10].

**Supplementary Methods 4: Subtype ambiguity reveals a continuous molecular spectrum with clinical implications**

To explore the continuous nature of subtype identity, we analyzed the relationship between ssGSEA-derived subtype scores (ΔZ) and model prediction ambiguity. Supplementary Figure S4a shows a clear linear relationship whereby samples closer to the decision boundary (ΔZ ≈ 0) exhibited higher ambiguity, while those with extreme scores showed low ambiguity. When modeled as a function of absolute distance from the boundary (see Supplementary Figure S4b), ambiguity distributions were curtailed, but interestingly, peak ambiguity centered slightly toward the basal spectrum rather than at the origin.

Analysis of ambiguity versus ssGSEA subtype scores (see Supplementary Figure S4c) revealed that samples with scores near 0.5 enrichment displayed the highest ambiguity, indicating genuine intermediate states. Importantly, no high-confidence samples (ΔZ > 1) demonstrated elevated ambiguity, confirming an inverse relationship between subtype confidence and prediction uncertainty.

GATA6 expression aligned strongly with the subtype axis (see Supplementary Figure S4d), with classical samples showing elevated GATA6 and basal-like samples showing reduced or absent expression. However, GATA6 expression versus ambiguity (see Supplementary Figure S4e) showed substantial overlap in the 0.6–0.8 ambiguity range across varying GATA6 levels.

Principal component analysis of samples using strong Moffitt genes, colored by ambiguity (see Supplementary Figure S4f), demonstrated that high-ambiguity samples clustered near the interface between classical and basal regions, while low-ambiguity samples formed tight, distinct clusters.

When analyzing variance in the top 10 Moffitt genes per subtype, higher variance corresponded to lower ambiguity, though most samples clustered within 5 variance units of the origin. High-variance samples at either tail were enriched for low-ambiguity cases, indicating that the subtype split is driven by variability in gene expression rather than consistent uniform enrichment.

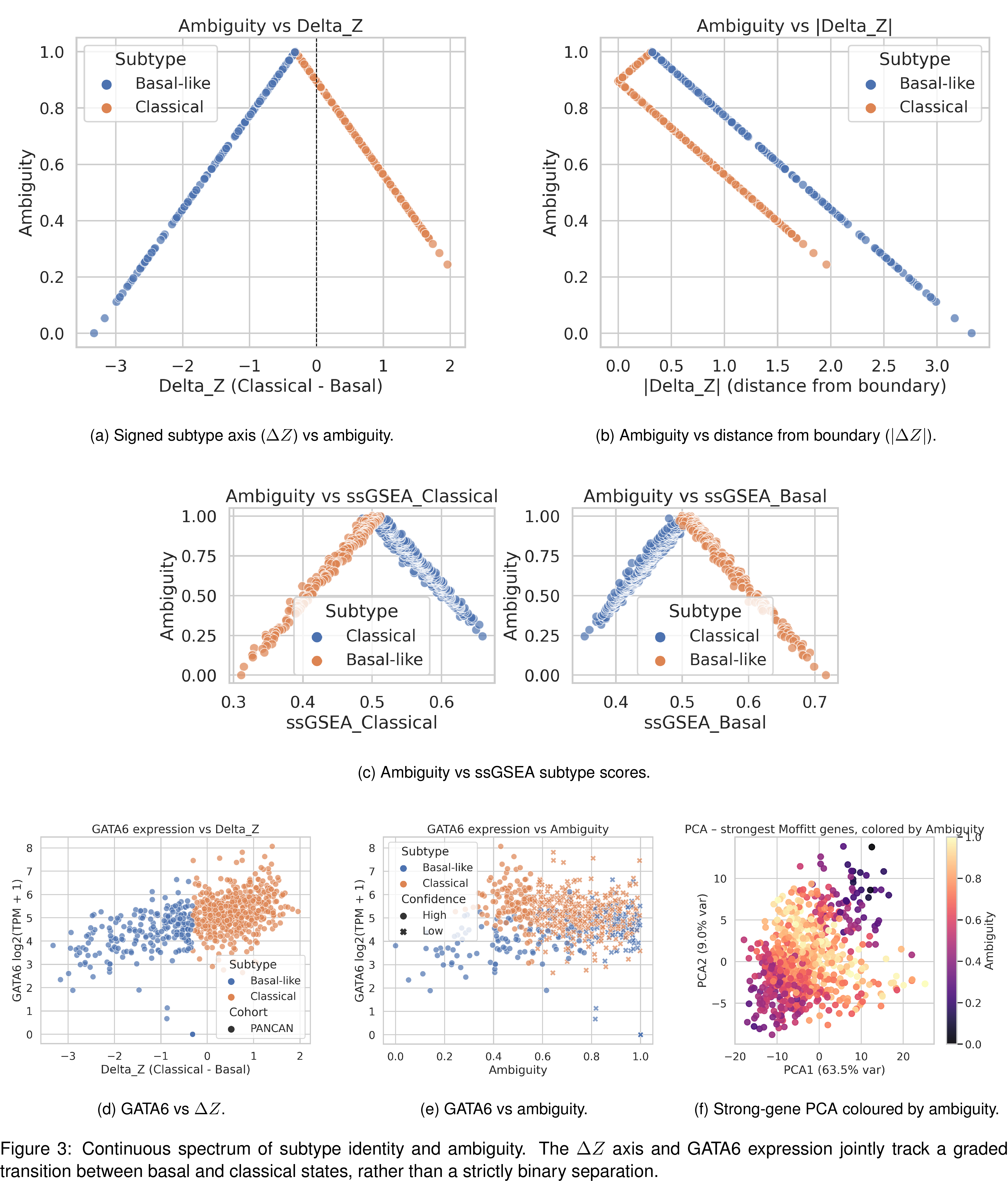

***Supplementary Figure S4:*** *Ambiguity score behavior, subtype separation, and biological correlates across PANCAN + TCGA samples. This figure characterizes the ambiguity score and its relationship to subtype-defining molecular features, ssGSEA signatures, GATA6 expression, and PCA structure.* ***(A)*** *Ambiguity vs. ΔZ (Classical – Basal score). Ambiguity is maximal near ΔZ = 0 (the subtype decision boundary) and decreases linearly as tumors diverge toward strongly Classical (positive ΔZ) or strongly Basal-like (negative ΔZ). Basal-like tumors populate negative ΔZ values, whereas Classical tumors populate the positive side.* ***(B)*** *Ambiguity vs. |ΔZ| (absolute distance from boundary). Absolute distance from the decision boundary is inversely correlated with ambiguity. Tumors farther from the boundary show exceptionally low ambiguity, whereas tumors near the center show high uncertainty.* ***(C)*** *Ambiguity vs. ssGSEA Classical enrichment. Classical tumors demonstrate higher ssGSEA enrichment for the Classical signature and correspondingly lower ambiguity. Basal-like tumors have reduced Classical ssGSEA scores and higher ambiguity near the transition zone.* ***(D)*** *Ambiguity vs. ssGSEA Basal enrichment. Basal-like tumors exhibit higher ssGSEA enrichment for the Basal signature, again with lower ambiguity as they move deeper into the Basal-like state. Classical tumors with weak Classical signatures show elevated ambiguity. Together, (C–D) validate that the ambiguity score aligns with biologically meaningful continuous variation, rather than classification noise.* ***(E)*** *GATA6 expression vs. ΔZ. GATA6 expression increases sharply with ΔZ, showing elevated levels in Classical tumors and low levels in Basal-like tumors. The gradient further supports GATA6 as a continuous indicator of subtype state.* ***(F)*** *GATA6 expression vs. ambiguity. Low-ambiguity tumors show clear subtype extremes: high GATA6 for Classical and low GATA6 for Basal-like. High-ambiguity tumors cluster near intermediate GATA6 expressions and include many low-confidence predictions.* ***(G)*** *PCA of strongest Moffitt genes, colored by ambiguity. Samples with high ambiguity cluster near the interface between Classical and Basal regions in PCA space. Clear subtype extremes exhibit low ambiguity and occupy opposite ends of the major expression gradient.*

**Supplementary Methods 5: DNA damage repair gene expression links molecular subtype to chemosensitivity**

Given the established association between DDR deficiency and platinum sensitivity, we examined the expression of key DDR genes across subtypes [11]. The Gene expressions of BRCA1/2, RAD51, ATM, PALB2, and CHEK1 were measured across all patients classified into the basal-like and classical category. Gene expression across all genes was summated and Z-scored across both subtypes and all cohorts into a stratified DDR expression score to visualize trends in gene expression within the basal and classical cohorts that indicate chemosensitivity. Median BRCA1 expression was similar between classical and basal-like cohorts (see Supplementary Figure S5a), but the classical cohort showed substantially greater spread, with the 25th percentile near zero expression. In contrast, basal-like tumors exhibited a tighter distribution centered at moderate expression. BRCA2 expression (see Supplementary Figure S5b) mirrored this pattern, though classical tumors displayed even lower expression at the 25th percentile.

Analysis of PALB2 (see Supplementary Figure S5c), RAD51 (see Supplementary Figure S5d), ATM (see Supplementary Figure S5e), and CHEK1 (see Supplementary Figure S5f) revealed similar trends. Classical tumors generally showed more stratified expression with greater variance, while basal-like tumors exhibited tighter distributions. Notably, median ATM expression was marginally higher in classical tumors, whereas median CHEK1 expression was lower in classical compared to basal-like tumors. The composite DDR score (see Supplementary Figure S5g), computed by aggregating expression across all DDR genes, showed that classical tumors had more stable, clustered expression profiles, whereas basal-like tumors displayed greater variability with numerous low-expression outliers.

Heatmap analysis ordered by ΔZ (see Supplementary Figure S5h) revealed that a large subset of samples expressed minimal or no DDR genes, distributed across both subtypes. When DDR score was plotted against ΔZ (see Supplementary Figure S5i), a linearly negative relationship emerged, though scattered with outliers predominantly in the basal-like cohort. This indicates that higher classical subtype confidence correlates with lower DDR expression on average. In high-confidence TCGA samples, multiple DNA damage response (DDR) genes exhibited significant subtype-specific expression differences independent of GATA6. CHEK1, RAD51, and BRCA2 showed higher expression in basal tumors compared with classical tumors (CHEK1: Welch p = 1.35 × 10⁻⁴, FDR = 5.41 × 10⁻⁴; RAD51: Mann–Whitney U p = 8.28 × 10⁻⁴, FDR = 2.21 × 10⁻³; BRCA2: Mann–Whitney U p = 1.80 × 10⁻², FDR = 3.59 × 10⁻²), whereas BRCA1, PALB2, and ATM were not significantly different after correction.

Modeling DDR score against prediction ambiguity (see Supplementary Figure S5j) showed no clear separation, though a substantial proportion of highly ambiguous samples clustered near zero DDR score, indicating no variability in DDR gene enrichment. A composite DDR score demonstrated a modest but significant shift between basal and classical tumors (Mann–Whitney U p = 3.29 × 10⁻²; FDR = 5.26 × 10⁻²). In survival analysis of 178 patients (93 events), high versus low DDR composite scores significantly stratified overall survival (log-rank p = 0.0031), in contrast to GATA6 expression, which was not prognostic, indicating that DDR activity captures prognostic information not explained by lineage alone.

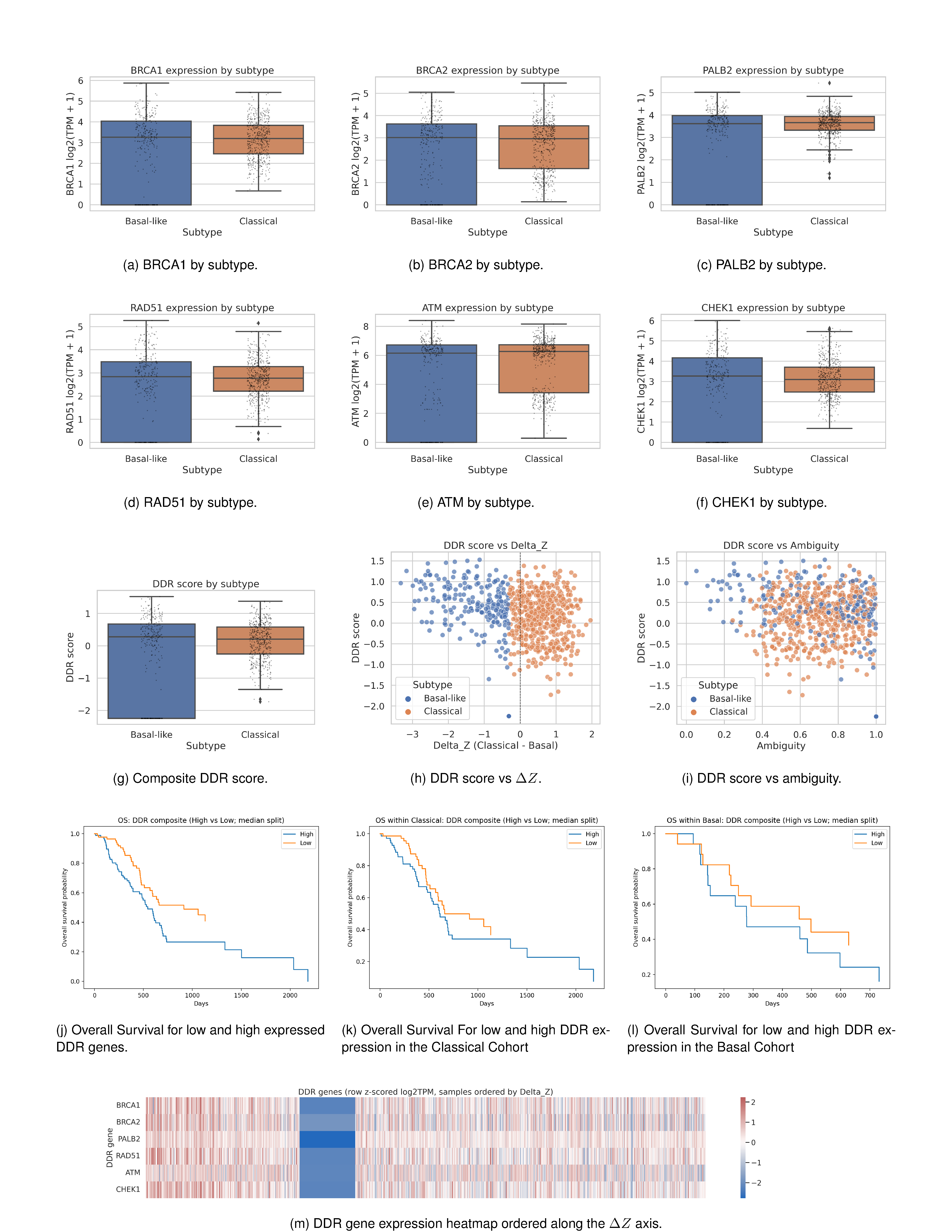

***Supplementary Figure S5:*** *DNA Damage Response (DDR) gene expression patterns across Basal-like and Classical tumors and their relationship to subtype continuum metrics. This figure examines expression of key homologous recombination and DDR genes across subtypes, integrates these genes into a composite DDR score, and evaluates how DDR activity varies across the Classical–Basal continuum. (****Top two rows:*** *Individual DDR gene expression by subtype) Boxplots display log2(TPM+1) expression for canonical homologous recombination repair (HRR) and DDR regulators across Basal-like and Classical tumors:* ***(A)*** *BRCA1,* ***(B)*** *BRCA2,* ***(C)*** *PALB2,* ***(D)*** *RAD51,* ***(E)*** *ATM,* ***(F)*** *CHEK1 Across all six genes, Classical tumors consistently show higher expression than Basal-like tumors. Basal-like tumors exhibit lower overall DDR component expression with greater heterogeneity, consistent with reduced HRR pathway activity. (****Third row:*** *Composite DDR score distributions and continuous relationships)* ***(G)*** *DDR score by subtype. A composite DDR score (computed as the mean z-scored expression of BRCA1, BRCA2, PALB2, RAD51, ATM, and CHEK1) is significantly lower in Basal-like tumors and higher in Classical tumors, mirroring individual gene-level patterns.* ***(H)*** *DDR score vs. ΔZ. DDR activity increases monotonically with ΔZ (Classical – Basal). Classical tumors (positive ΔZ) show clear enrichment for DDR signaling, whereas Basal-like tumors (negative ΔZ) cluster toward reduced DDR activity.* ***(I)*** *DDR score vs. Ambiguity. Higher ambiguity is associated with lower DDR activity, particularly among Basal-like tumors. Classical tumors with strong DDR signaling display low ambiguity, further supporting the biological interpretability of the ambiguity metric.* ***(J–L)*** *Kaplan–Meier overall survival analyses stratified by high versus low DNA damage response (DDR) composite score (median split), shown for the full cohort (j), the classical subtype (k), and the basal subtype (l). Survival associations differ by subtype, with significant stratification observed in the full cohort and classical tumors, but not in basal tumors.* ***(M)*** *Heatmap of DDR genes (row-z-scored log2TPM), samples ordered by ΔZ. Samples are arrayed from strongly Basal-like (left) to strongly Classical (right). DDR gene expression shows a coordinated gradient: Basal-like tumors exhibit systematic downregulation (blue), while Classical tumors demonstrate cohesive upregulation (red). This provides visual confirmation of DDR pathway strengthening along the Classical direction.*

**Supplementary Methods 6: Gene ontology and pathway analysis reveal distinct functional programs**

To assess whether the model captured biologically functional rather than purely statistical features, we examined the biological programs represented by subtype-associated genes using Gene Ontology (GO) and KEGG pathway enrichment analyses of the top 25 classical and basal-like genes [12, 13]. GO Biological Process enrichment (see Supplementary Figure S6a, c–d) showed that classical-associated genes were predominantly linked to digestive system processes, digestion, and epithelial tissue development. These enrichments were driven by genes such as REG4, TFF1/2/3, CEACAM5, and LGALS4, which are established mediators of mucin secretion, epithelial barrier integrity, and pancreatic ductal function [54]. Additional enrichment was observed for epithelial structural maintenance and gastrointestinal tract development, with weaker associations involving metabolic regulation and anoikis resistance.

In contrast, basal-like–associated genes exhibited strong enrichment for tissue development, epidermal differentiation, and wound-associated programs. Notably, a substantial fraction of genes annotated to tissue development also mapped to wound healing, keratinization, and extracellular matrix remodeling, consistent with the regenerative, mesenchymal, and stress-responsive phenotypes characteristic of basal-like tumors [55].

Consistent with these findings, KEGG pathway analysis (see Supplementary Figure S6b, e–f) identified significant enrichment of cornified envelope formation within the basal-like gene set, involving eight highly significant genes, a pathway closely associated with squamous differentiation and wound repair [55]. In comparison, only a single classical-associated gene showed weak enrichment for this pathway. These molecular programs closely parallel the squamous histomorphology observed in basal-like pancreatic ductal adenocarcinoma [14], demonstrating that the model recovers subtype-specific biological processes rather than relying on uninterpretable image correlations.

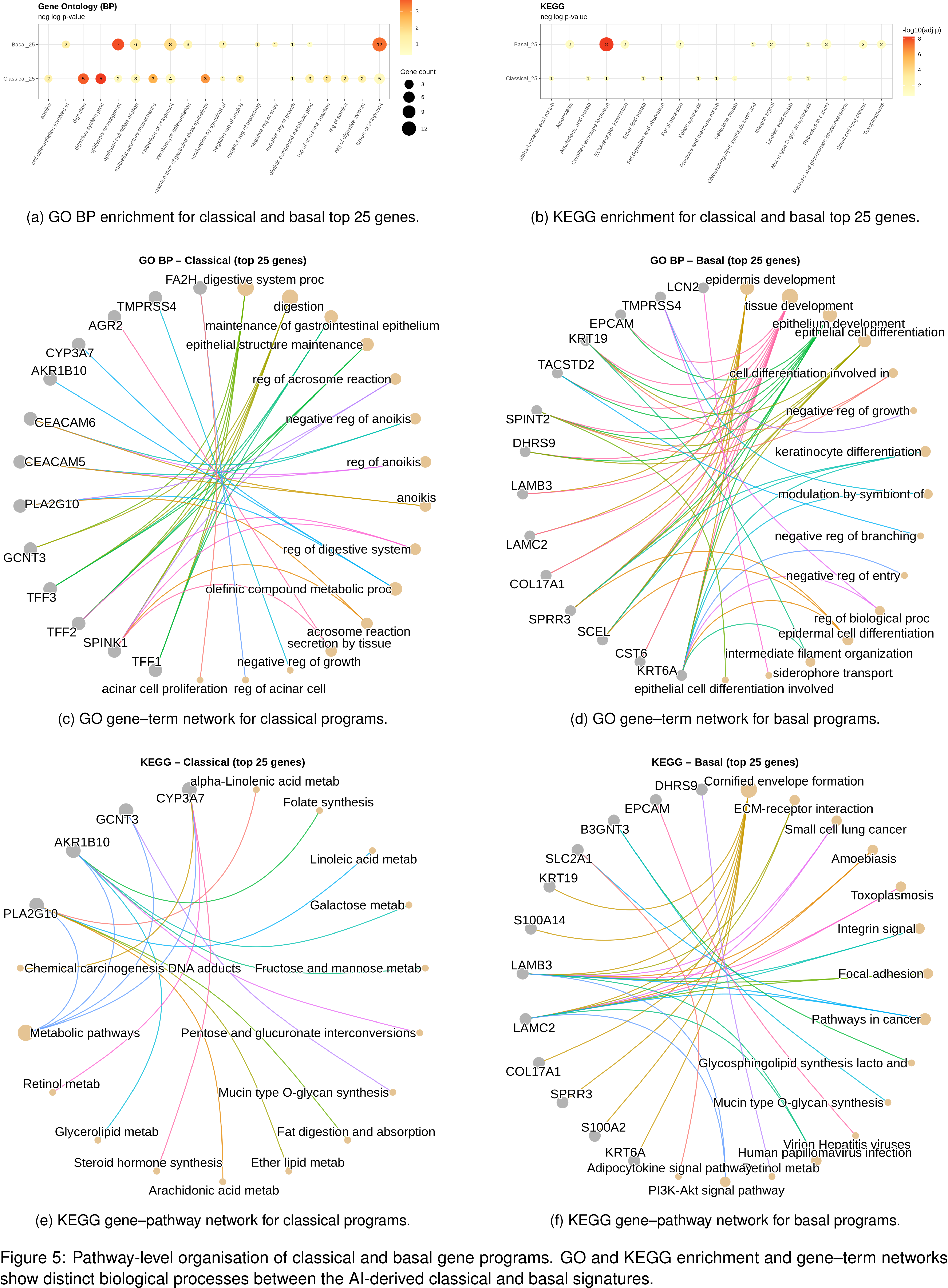

***Supplementary Figure S6:*** *Functional enrichment analysis of Classical and Basal-like subtype–specific genes reveal distinct biological programs and pathway specializations. This figure summarizes Gene Ontology (GO) and KEGG enrichment patterns for the top differentially expressed genes defining the Classical and Basal-like pancreatic cancer subtypes. Bubble plots provide high-level enriched terms, while chord diagrams map individual genes to their associated pathways, highlighting subtype-specific biological roles.* ***(A)*** *Gene Ontology Biological Process (GO BP) enrichment. Classical subtype genes are enriched for digestive, metabolic, acinar-cell, and epithelial maintenance processes. Basal-like genes show enrichment in keratinocyte differentiation, epithelial development, epidermal structure, and stress- or injury-related processes. Bubble color represents –log10 adjusted p-value; size represents gene count.* ***(B)*** *KEGG pathway enrichment. Classical subtype genes preferentially map to lipid metabolism, xenobiotic metabolism, steroid synthesis, and other homeostatic epithelial functions. Basal-like subtype genes show strong enrichment in ECM–receptor interaction, focal adhesion, PI3K–Akt signaling, pathways in cancer, and infection-related KEGG pathways, consistent with a more mesenchymal and stress-associated transcriptional state.* ***(C)*** *Classical subtype – top 25 GO BP genes. Chord diagram links Classical-specific genes to enrich biological processes such as digestive system function, epithelial structure maintenance, acinar cell regulation, and olefinic compound metabolism. Prominent genes include CEACAM6, TMPRSS4, AGR2, CYP3A7, and AKR1B10, emphasizing classical epithelial identity and metabolic specialization.* ***(D)*** *Basal-like subtype – top 25 GO BP genes. Basal-like genes connect to differentiation programs (epidermal, keratinocyte, epithelial), negative regulation of growth, branching morphogenesis, and modulation of biological processes. Key drivers include KRT6A, KRT19, LCN2, LAMC2, and SPRR3, reflecting a wound-like, regenerative, and injury-responsive phenotype.* ***(E)*** *Classical subtype – top 25 KEGG genes. Classical genes map to lipid metabolism (arachidonic acid, alpha-linolenic acid), carbohydrate metabolism (galactose, fructose/mannose), retinol metabolism, steroid hormone synthesis, and chemical carcinogenesis pathways. These enrichments match a differentiated, metabolically active epithelial lineage.* ***(F)*** *Basal-like subtype – top 25 KEGG genes. Basal-like genes connect to ECM–receptor interaction, focal adhesion, integrin signaling, PI3K–Akt signaling, pathways in cancer, and viral/infection-related pathways. Genes such as LAMC2, COL17A1, DHRS9, S100A2, and EPCAM highlight extracellular matrix remodeling, motility, and stress-response biology.*
